# Impact of Pre-Existing Adenovirus Immunity on Vaccine Immunity Induced by ChAdOx1 nCoV-19 in Immunodeficient Patients

**DOI:** 10.64898/2026.05.27.26354282

**Authors:** Ernest T. Aguinam, Andrew CY Chan, George W Carnell, Angalee Nadesalingam, Benedikt Asbach, Javier Castillo-Olivares, Ralf Wagner, Barbara Blacklaws, Helen Baxendale, Jonathan L Heeney

## Abstract

**Introduction:** Adenoviral vectors such as chimpanzee ChAdOx1 were selected for COVID-19 vaccines due to their low seroprevalence in humans, minimizing the impact of neutralising anti-vector immunity that could attenuate vaccine responses. However, the influence of pre-existing adenoviral immunity on vaccine response remains incompletely understood. We have previously shown that SARS-CoV-2 spike-specific T cells were enhanced in ChAdOx1 nCoV-19 vaccinated immunodeficient patients compared to mRNA-based BNT162b2. Here, we assess immune cross-reactivity between ChAdOx1 and human adenovirus 5 (HuAd5), and test the hypothesis that in antibody-deficient individuals, cross-neutralisation may be impaired, allowing bystander enhancement of SARS-CoV-2 spike-specific T cell responses following ChAdOx1 nCoV-19 vaccination.

**Methods:** We studied healthy healthcare workers (HCWs) and immunodeficient patients (IDPs) who received homologous ChAdOx1 nCoV-19 or BNT162b2 vaccines. HCWs samples were collected pre-vaccination and 4-6 weeks after the second dose, while IDP samples were obtained 4-6 weeks after the second dose. Serum anti-HuAd5 hexon IgG was quantified using a Luminex multiplex assay, and neutralizing antibodies were assessed using a replication-deficient HuAd5-GFP virus neutralization assay with flow cytometry readout. *Ex vivo* ELISpot and flow cytometry assays were used to measure T cell responses to HuAd5 hexon. These data were compared with previously published ChAdOx1 nCoV-19 vaccine responses in the same cohorts.

**Results:** HuAd5 hexon-binding IgG titres were significantly higher in ChAdOx1 nCoV-19 compared to BNT162b2 vaccine recipients in both HCWs (p = 0.0043) and IDPs (p = 0.0328). Within ChAdOx1 nCoV-19 vaccine group, titres were lower in IDPs than HCWs (p = 0.0015) but not within the BNT162b2 group (p = 0.1261). HuAd5 neutralisation titres did not differ between cohorts or vaccine groups. In ChAdOx1 nCoV-19 vaccinated IDPs and HCWs, there was a significant negative correlation between HuAd5 hexon IgG titres and SARS-CoV-2 spike-specific T cell responses. Similarly, HuAd5 neutralisation titres showed an inverse correlation with spike-specific T cell responses in ChAdOx1 nCoV-19 vaccinated IDPs and HCWs. ChAdOx1 nCoV-19 vaccination induced significantly higher frequencies of HuAd5 hexon-reactive T cells compared with BNT162b2 vaccination in IDPs (p < 0.0001), consistent with cross-reactive adenoviral T cell responses. In IDPs, HuAd5 hexon-specific T cell frequencies positively correlated with SARS-CoV-2 spike-specific T cell responses following ChAdOx1 nCoV-19 vaccination but not following BNT162b2 vaccination. Functional profiling in ChAdOx1 nCoV-19 vaccinated IDPs demonstrated expansion of HuAd5 hexon-specific CD4⁺IFN-γ⁺TNFα⁺ T cells in high SARS-CoV-2 spike responders (p = 0.0002) compared to low responders, and the frequency of these cells strongly correlated with spike-specific T cell response.

**Discussion:** ChAdOx1 nCoV-19 has been associated with stronger T cell responses than BNT162b2 in certain populations, including immunodeficient and elderly individuals. While this has been attributed to antigen persistence and innate adjuvant effects, our findings support a mechanism whereby heterologous pre-existing adenovirus immunity modulates vaccine-induced responses. Specifically, cross-reactive HuAd5-specific T cells may enhance spike-specific T cell responses via bystander enhancement, while cross-reactive binding antibodies may exert opposing effects. An implication of this study is that vaccine protocols could incorporate therapies that suppress vector-specific or cross-reactive antibodies while preserving T cell responses especially in cases where T cell-specific responses are most desirable. Also, safe vector-based vaccines can be developed for patient groups with predominant antibody deficiency. Targeted vaccination strategy could be implemented for clinical cohorts based on immune competence.

## INTRODUCTION

Viral vectors have been extensively used in vaccine formulations to deliver transgenes encoding immunogens of interest for infectious disease prevention and cancer therapy ^1^. The COVID-19 pandemic response saw the licensing of several adenovirus-based vaccines encoding the SARS-CoV-2 spike protein ^2,3^. One concern about the use of viral vector technology has been that anti-vector immunity (specific or cross-reactive) acquired from previous infections and/or vaccinations may dampen the immune response to the transgene product of interest. This is of particular concern with regard to some human adenoviruses that are highly prevalent in the population such as human adenovirus 5 (HuAd5) where the seroprevalence is reported to range between 40 – 90 % ^4^. Studies have reported a reduced cellular immune response to adenovirus-vectored vaccine inserts when there is a pre-existing immune response ^5–8^. To circumvent this, recent adenovirus-based vaccine designs have used less prevalent and non-human adenoviruses ^3,9,10^. The ChAdOx1 vector, derived from chimpanzee adenovirus Y25 (ChAdY25), was selected for further development as a vector for human vaccines and therapeutics based on a low seroprevalence in the human population, and its ability to induce comparable cellular immunogenicity to other adenovirus vectors ^11^. Recent evidence of T cell cross-reactivity between human adenovirus-specific T cells and ChAdOx1 has been presented ^12^. T cell and antibody cross-reactivity has also been identified between other heterologous adenoviruses ^13–15^, but the extent to which these heterologous pre-existing vector responses, cell-mediated and humoral, may affect the response to a new antigen expressed by the adenovirus vaccine is not fully understood.

Optimisation of immunisation strategies for vulnerable patients such as those with immune deficiencies, is a particular challenge ^16^. Notably, antibody responses are more commonly impaired, whilst cell-mediated responses may be intact. We and others have previously reported a significantly enhanced induction of SARS-CoV-2 spike T cell responses by ChAdOx1 nCoV-19, compared to the mRNA-based BNT162b2 vaccine in immunodeficient or elderly individuals ^17–21^. In our study, the sustained difference after two doses of the vaccines was unique to an immunodeficient cohort as compared with healthy controls. Most of these patients (65 %) had a predominant antibody deficiency and were receiving regular immunoglobulin replacement therapy as part of their clinical management. To delineate possible mechanisms underlying the differences in vaccine-induced T cell responses, we hypothesized that antibody-deficient individuals generate enhanced activation of adenovirus-specific memory T cells that cross-recognise conserved epitopes in the ChAdOx1 nCoV-19 vaccine vector due to reduced antibody-mediated neutralisation of vector and bystander enhancement of SARS-CoV-2 reactive T cells. In contrast, healthy individuals may generate heterologous (cross-reactive) and homologous adenoviral vector antibody responses that limit the induction of anti-vector T cell responses and subsequent bystander activation of SARS-CoV-2 specific T cells through early inhibition of vector antigen expression.

Bystander enhancement, a phenomenon in which T cells are activated non-specifically by inflammatory mediators, cytokines and ligands produced by other immune cells, has been described in the context of viral infections and autoimmune diseases ^22–24^. There have also been reports of bystander activation of naïve and unrelated memory CD4^+^ T cells ^25,26^ and CD8^+^ T cells ^27^. Essentially, naïve T cells may undergo activation, and antigen-specific effector or memory T cells can be activated more efficiently in an antigen-independent manner when stimulated by sufficient levels of cytokines. It is well known that three distinct signals are required in a typical immune response for T cell activation and expansion namely, T cell receptor recognition of antigen bound to MHC molecules, the interaction of co-stimulatory molecules and cytokine signalling ^28^. This third signal from cytokines is important for the expansion of recently activated cells which typically upregulate their expression of cytokine receptors thus lowering their threshold for cytokine activation in comparison to naïve cells^23,28^. The expansion of the newly activated clones of SARS-CoV-2 specific T cells in the predominantly antibody-deficient cohort^17^, may have been enhanced by heightened levels of cytokines produced by anti-human adenovirus T cells that were boosted by cross-reactivity to the ChAdOx1 vector.

Enhanced T cell responses following ChAdOx1 vaccination have been attributed to vector-related adjuvant effects and differences in antigen presentation ^18,29^. Adenoviral vectors may promote prolonged antigen persistence due to delayed trafficking to draining lymph nodes and transduction of non-immune cells such as fibroblasts, resulting in sustained low-level antigen expression and potentially inflated memory responses ^30,31,29^. In contrast, mRNA vaccines such as BNT162b2 exhibit rapid antigen clearance at the injection site, although spike protein can persist transiently in the circulation and mRNA can remain detectable in innate immune cells for several days ^32^. These differences in antigen kinetics and cellular targeting likely contribute to the distinct immune profiles observed between vaccine platforms.

Innate immune signalling further shapes these responses. Replication-deficient adenoviral vectors efficiently transduce dendritic cells and induce type I interferon production, which promotes T cell activation and differentiation ^33^. Pre-pandemic PBMC studies have shown that ChAdOx1 stimulation can activate memory CD4⁺ T cells and induce a pro-inflammatory cytokine profile ^12^. However, mRNA vaccines also elicit strong innate immune activation, including recruitment and activation of several innate immune cells accompanied by production of type I interferons and other cytokines ^32,34^.

But differences in antigen presentation and innate immune activation do not fully explain the enhanced spike-specific T cell responses observed in ChAdOx1 nCoV-19 vaccinated IDPs. The absence of a similar differential response in HCWs ^17^ suggests that host factors specific to the IDP cohort contribute to this effect.

To test this hypothesis that ChAdOx1-based vaccine response is modulated by pre-existing cross-reactive human adenovirus immune memory, we used available samples from the previous cohort study ^17^ who had received double homologous doses of either ChAdOx1 nCoV-19 or BNT162b2. We assessed antibody and T cell responses against HuAd5 and their association with the immune responses to the vaccine-encoded spike protein as previously reported ^17^. HuAd5 was chosen as a representative human adenovirus due to its high prevalence in humans ^4^. Sequence alignment of the hexon and penton proteins of ChAdY25 and HuAd5 (NCBI, protein blast) showed a 77.2 % similarity for hexon and 70.7 % similarity for penton base proteins (Accession numbers: ChAdY25 hexon = YP_006272963.1, HuAd5 hexon = BAG48782.1; ChAdY25 penton = YP_006272958, HuAd5 penton base = AAA42519). Here, we present experimental evidence of antibody and T cell cross-reactivity between HuAd5 and ChAdOx1 vector and supporting evidence of a positive effect of pre-existing human adenovirus memory T cell on ChAdOx1 nCoV-19 vaccine T cell responses in predominantly antibody-deficient individuals.

## RESULTS

### Enhanced HuAd5 hexon IgG responses in ChAdOx1 nCoV-19 vaccine recipients

To understand the interaction between ChAdOx1 vaccine vector and the pre-existing immunity to closely related HuAd5, we assessed HuAd5 hexon-binding IgG titres in IDPs and HCWs after double homologous doses of ChAdOx1 nCoV-19 vaccine (n = 57 and n = 37 respectively). As controls, we also tested for HuAd5 hexon IgG in BNT162b2 vaccinated IDPs and HCWs (n = 38 and n = 42 respectively). We observed that ChAdOx1 nCoV-19 vaccinated HCWs had higher titres of HuAd5 hexon-binding IgG compared to BNT162b2 vaccinated HCWs (p = 0.0043, Fig 2A).

**Figure 1:**
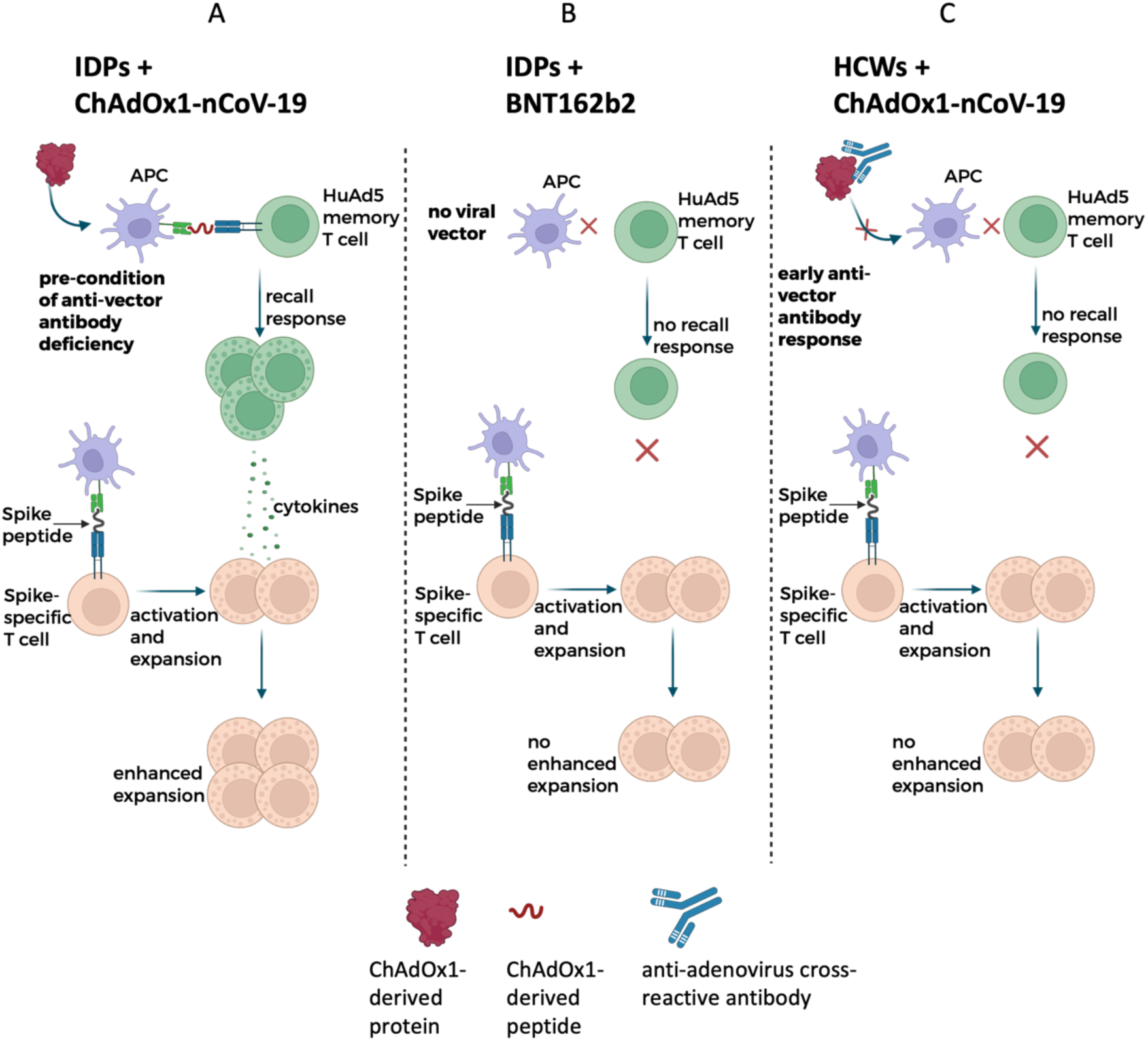
Model of bystander enhancement of ChAdOx1 nCoV-19 responses in immunodeficient patients (IDPs) **A)** In antibody deficient IDPs receiving ChAdOx1 nCoV-19, cross-reactive HuAd5 specific T cells are recalled, expand, and produce cytokines that promote enhanced spike-specific T cell expansion. **B)** In IDPs vaccinated with BNT162b2, no HuAd5 specific T cell recall occurs, so additional cytokine support and bystander spike T cell expansion are absent. **C)** In healthy controls (HCWs) receiving ChAdOx1 nCoV-19, anti-adenovirus antibodies limit vector expression and prevent HuAd5 specific T cell recall, resulting in no enhanced spike-specific T cell expansion.

**Figure 2.**
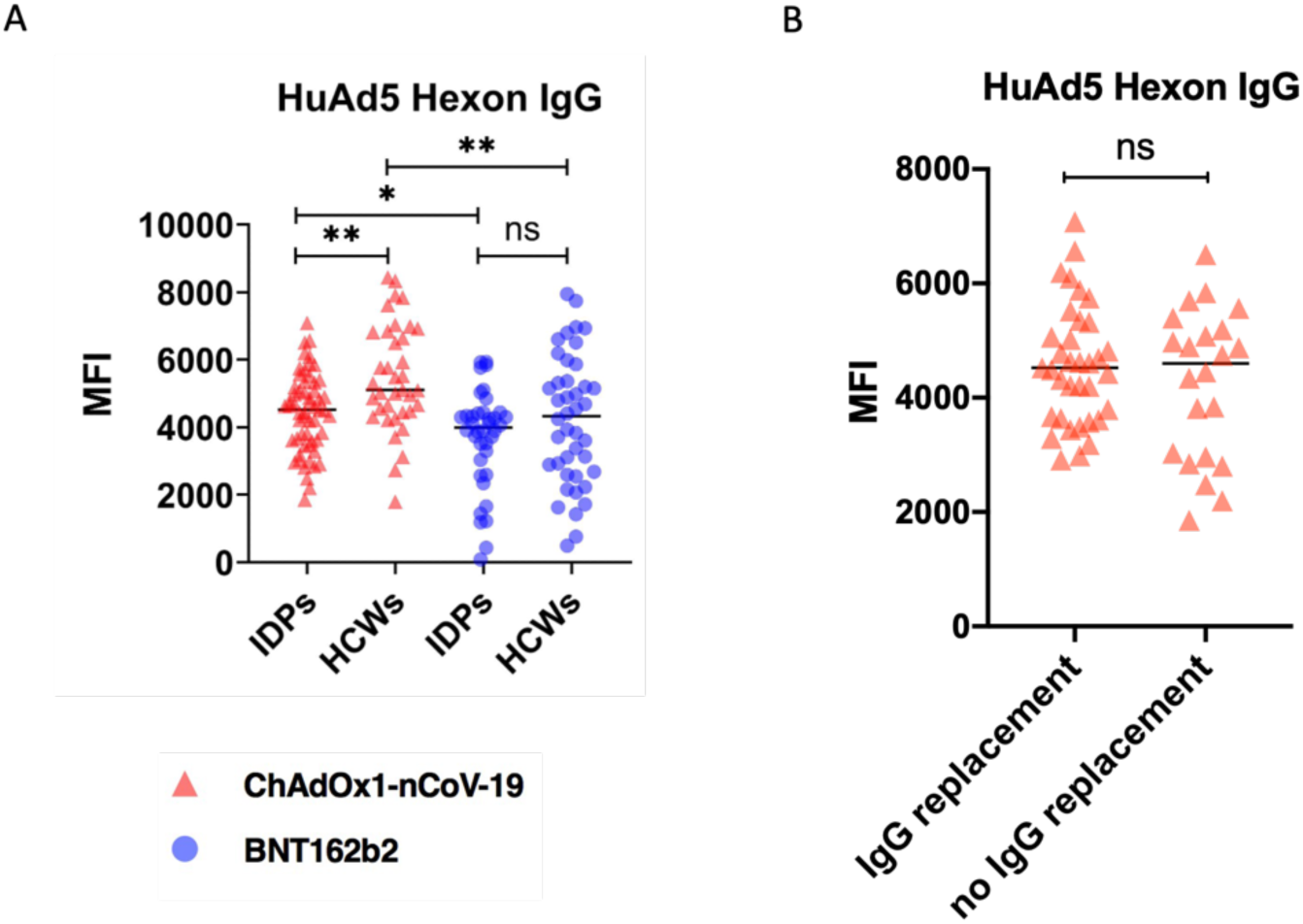
HuAd5 Hexon-specific IgG Antibody Responses. Serum anti-hexon IgG titres in HCWs and IDPs were measured using a Luminex assay. **A)** Anti-hexon IgG titres in IDPs and HCWs vaccinated with either ChAdOx1 nCoV19 (n = 57 and n = 37 respectively) or BNT162b2 (n = 38 and n = 42 respectively). **B)** Comparison of anti-hexon IgG titres based on IgG replacement history (treatment: n = 35; no treatment: n = 22). Data points represent the mean of duplicate wells minus negative controls. Statistical differences were determined using non-parametric Mann Whitney test. * is p< 0.05, ** is p < 0.01, ns = not significant. ChAdOx1 nCoV-19 group (IDPs: n = 57 and HCWs: n = 37). BNT162b2 group (IDPs: n = 38 and HCWs: n = 42)

Similarly, ChAdOx1 nCoV-19 vaccinated IDPs had higher titres than their BNT162b2 vaccinated counterpart although to a lesser extent than in the HCWs (p=0.0328, Fig 2A). Inter-cohort comparison within vaccine groups showed that ChAdOx1 nCoV-19 vaccinated HCWs had significantly higher titres than IDPs (p = 0.0015) whereas there was no difference in titres between the BNT162b2 vaccinated IDPs and HCWs (p = 0.2161) (Fig 2A). These results suggest that ChAdOx1 nCoV-19 vaccination induced HuAd5 hexon IgG response in both HCWs and IDPs.

As a majority of the IDPs received IgG replacement treatment due to severe antibody deficiency, we analysed data to determine whether there were any differences in antibody levels in ChAdOx1 nCoV-19 vaccinees based on IgG replacement treatment history. We observed no differences in HuAd5 IgG titres between those receiving treatment (n = 35) and those not receiving treatment (n = 22) (Fig 2B).

### HuAd5 neutralisation titres are similar across cohorts and vaccine groups

To further characterise functional anti-HuAd5 antibodies, we assessed serum HuAd5 neutralisation. A549 cells expressing GFP upon HuAd5-GFP infection were observed by fluorescent microscopy and enumerated by flow cytometry, with representative slides and gating strategy shown in Figure 3A & B. Unlike with the HuAd5 hexon IgG levels, there was no difference in the HuAd5 neutralisation titres between HCWs and IDPs or between vaccine groups (Fig 3C). There was also no difference in neutralisation titres based on IgG replacement therapy (Fig 3D). We observed a wide range of responses across the main sub-cohorts but responses in IDPs receiving IgG replacement therapy clustered within a narrower range.

**Figure 3:**
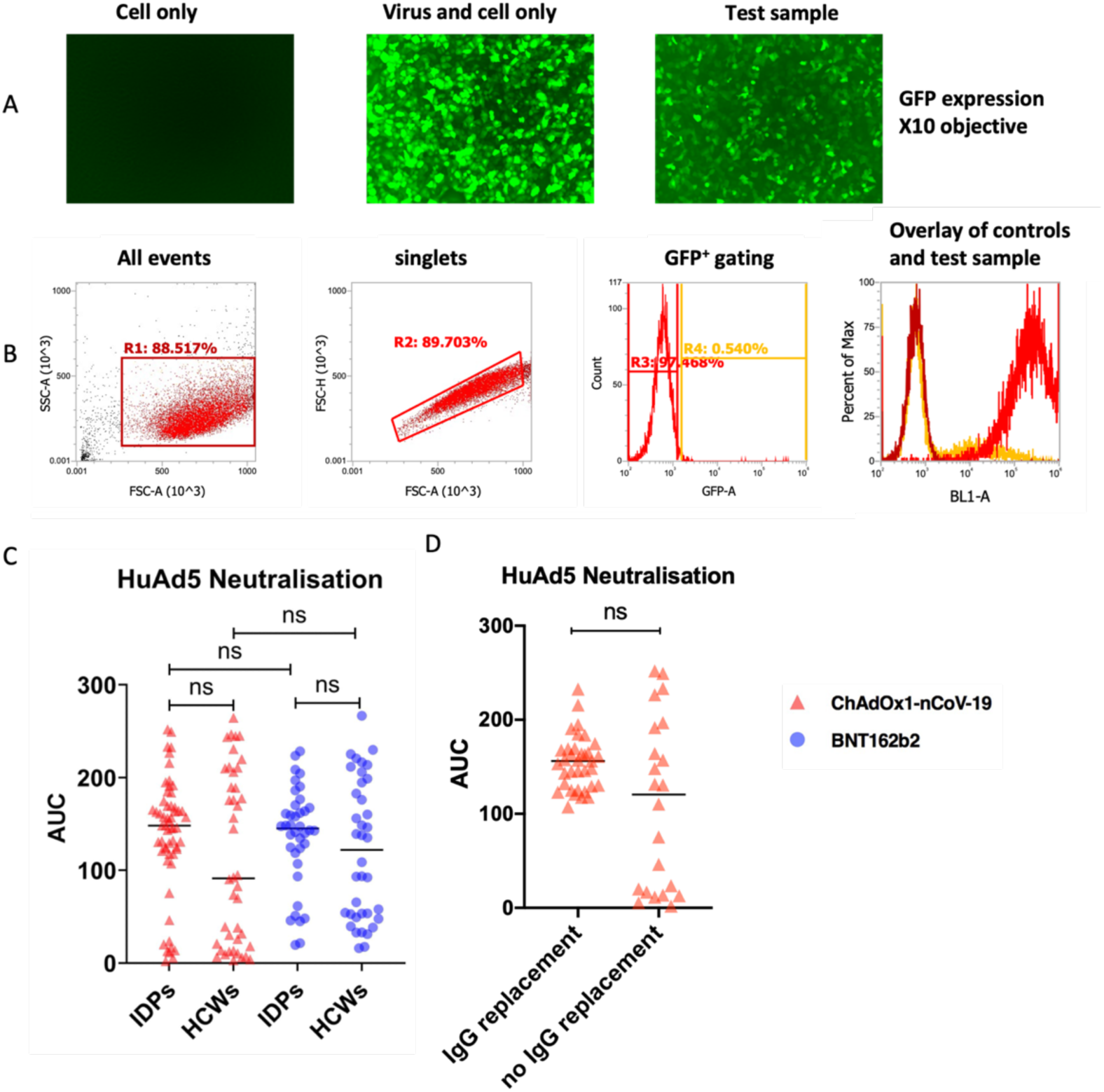
HuAd5-GFP Neutralizing Antibody Responses. Serum HuAd5 neutralising antibody titres were assessed using a A549 cell-line-based HuAd5-GFP neutralisation assay. **A)** Representative slides demonstrating GFP expression in A549 cells as observed under the fluorescent microscope **B)** Graphs showing flow cytometry gating for detection of GFP^+^ cells. **C)** HuAd5-GFP neutralizing antibody titre in HCWs and IDPs. N = 57 and 37 for IDPs and HCWs respectively for ChAdOx1 nCoV-19 group and, for BNT162b2 group, n = 38 and 38 for IDPs and HCWs respectively. **D)** Comparison of HuAd5 neutralisation based on IgG replacement history (treatment: n = 35; no treatment: n = 22). Neutralising antibody responses are expressed as Area Under the Curve (AUC) calculated from 5 point 5-fold serial dilution of serum samples. Group comparisons were performed using Mann-Whitney tests. ns = not significant.

### HuAd5 hexon-specific IgG titres correlated inversely with SARS-CoV-2 spike-specific T cell frequency in ChAdOx1 nCoV-19 vaccinated IDPs and HCWs

To understand the relationship between anti-HuAd5 antibodies and ChAdOx1 nCoV-19 vaccine T cell responses, and in particular, to investigate our hypothesis that cross-reactive anti-vector antibody can modulate ChAdOx1 nCoV-19 vaccine-induced spike-specific T cells, we performed correlation analysis between HuAd5 hexon IgG titres and our previously reported SARS-CoV-2 spike T cell responses^17^. We aimed to determine whether low-titre hexon-binding IgG is associated with enhanced spike-specific T cell responses seen in immunodeficient patients following ChAdOx1 nCoV-19 vaccination. Indeed, we observed significant inverse correlation between HuAd5 hexon IgG antibody levels and SARS-CoV-2 spike-specific T cell responses in ChAdOx1 nCoV-19 vaccinated IDPs and HCWs although the correlation was weaker in IDPs (IDPs: Spearman r = -0.3229, p = 0.0143; Fig 4A and HCWs: Spearman r = -0.5699, p = 0.007; Fig 4B). Similar analysis was performed for HuAd5 neutralising antibody titres. Negative correlation between HuAd5 neutralisation and SARS-CoV-2 spike-specific T cell responses was also observed for IDPs (Spearman’s r = -0.2917, p = 0.0467, Fig 4C), and HCWs (Spearman’s r = -0.5618, p = 0.0247, Fig 4D). HuAd5 hexon IgG binding titres correlated positively with HuAd5 neutralisation titres in both cohorts: IDPs (Spearman’s r = 0.3303, p

**Figure 4.**
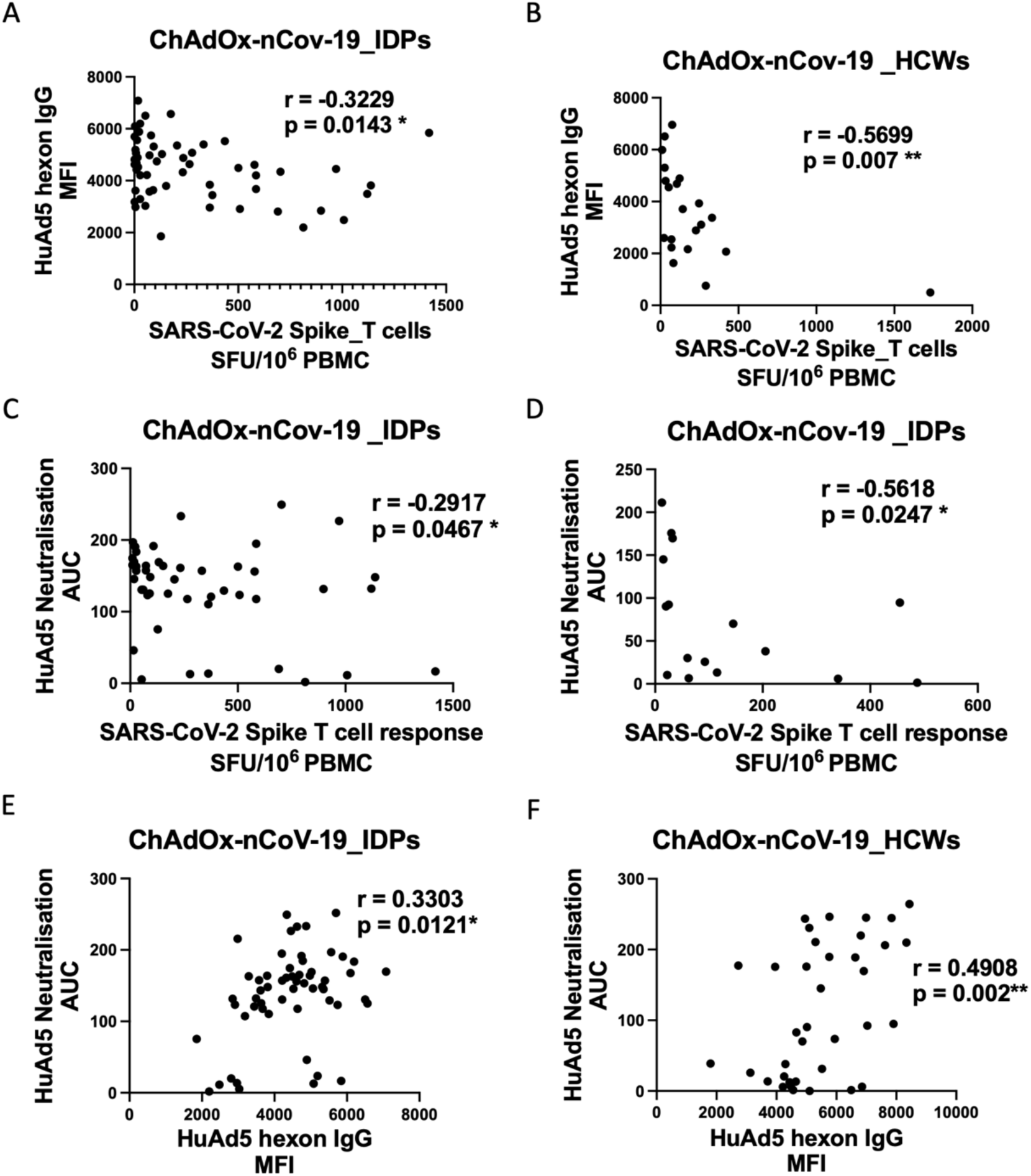
Relationship between HuAd5 antibody responses and ChAdOx1 nCoV-19 induced spike T cell responses. A & B) Correlation between HuAd5 hexon IgG titres and spike T cell response in **A)** IDPs and **B)** HCWs. C & D) Correlation between HuAd5 neutralization and spike T cell response in **C)** IDPs and **D)** HCWs. E & F) Correlation between HuAd5 hexon IgG titres and HuAd5 neutralization in **E)** IDPs and **F)** HCWs.

= 0.0121, Fig 4E) and HCW (Spearman’s r = 0.4908, p = 0.002, Fig 4F).

### Frequency of HuAd5 hexon-specific T cells was higher in ChAdOx1 nCoV-19 compared to BNT162b2 vaccinated IDPs

An important premise of our hypothesis of bystander enhancement by pre-existing human adenovirus-specific T cells is the occurrence of significant T cell cross-reactivity between human adenoviruses and the ChAdOx1 vector. Initial testing by IFN-γ ELISpot of 15 paired samples from healthy controls pre- and 4 – 6 weeks post-first vaccination, showed there was no significant difference in HuAd5 hexon-specific T cells between ChAdOx1 nCoV-19 and BNT162b2 groups before vaccination (p=0.0568, Supplementary Fig 1A), however, ChAdOx1 nCoV-19 vaccinees had significantly higher frequencies of HuAd5 hexon-specific T cells compared to BNT162B2 vaccinees after first vaccination (p=0.0068, Supplementary Fig 1B), suggesting that the ChAdOx1 vector induces cross-reactive HuAd5 T cell responses. We had previously demonstrated that in this cohort of IDPs, the T cell responses to positive control peptides representing commonly encountered antigens were similar between the two vaccine cohorts^17^. When we assessed HuAd5 hexon-specific T cell responses in the IDPs, we observed significantly higher frequencies of reactive T cells in the ChAdOx1 nCoV-19 vaccinees (n = 46) compared to the BNT162b2 vaccinees (n = 25) (p>0.0001, Fig 5A).

**Figure 5:**
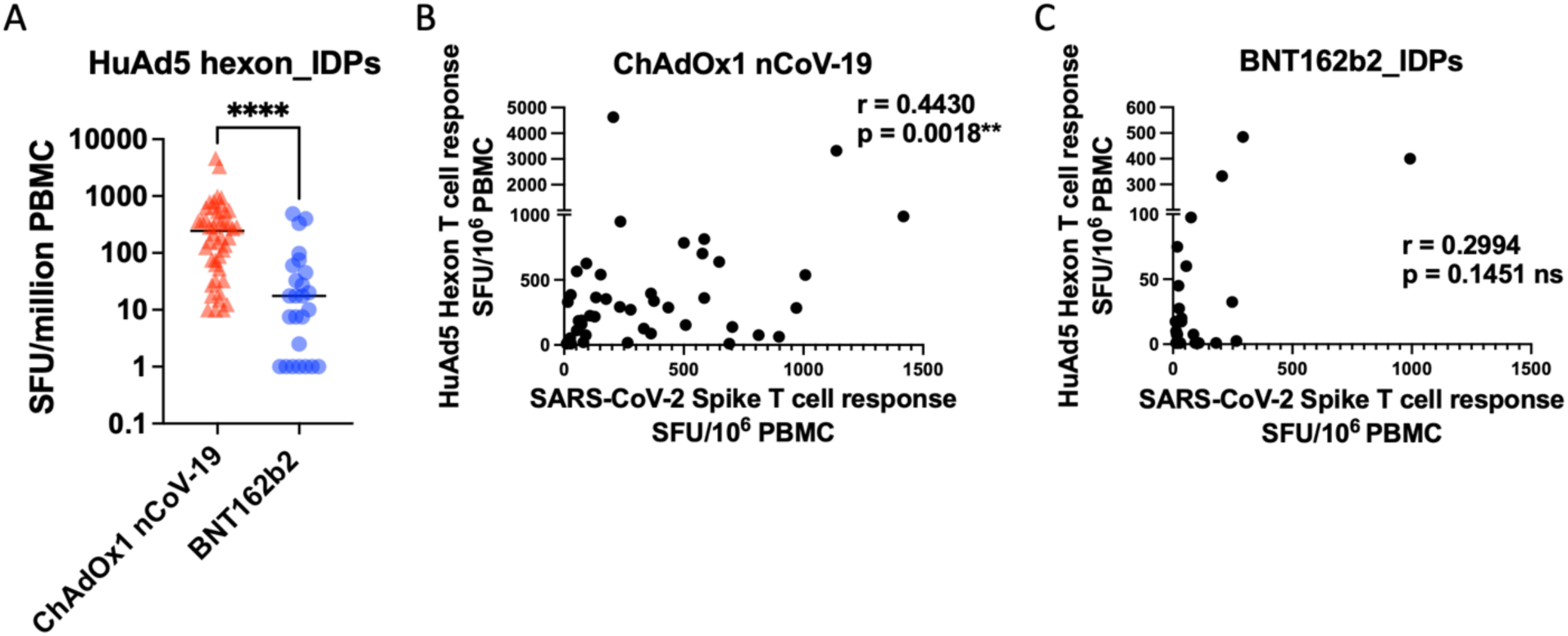
HuAd5 hexon-specific T Cell Responses in IDPs. PBMC samples from IDPs were tested using ELISpot for HuAd5 hexon-specific T cell responses. **A)** Comparison of HuAd5 hexon-specific T cell responses between ChAdOx1 nCoV-19 (n = 46) and BNT162b2 (n = 25) vaccinated IDPs. B & C) Correlation between HuAd5 hexon-specific T cell responses and the previously reported SARS-CoV-2 spike-specific T cells responses in **B)** ChAdOx1 nCoV-19 vaccinated and **C)** BNT162b2 vaccinated IDPs Significant difference was determined by non-parametric Mann Whitney test: **** is p < 0.0001. Spearman’s r and p values are shown within the correlation graph.

To determine whether high HuAd5 hexon T cell responses were associated with the enhanced spike-specific T cell responses in ChAdOx1 nCoV-19 vaccinees, we performed a correlation analysis between HuAd5 hexon and the previously reported SARS-CoV-2 spike T cell responses^17^. We observed a significant positive correlation in ChAdOx1 nCoV-19 vaccinated IDPs (Spearman’s r = 0.4412, p = 0.0021, Fig 5B) but not in BNT162b2 vaccinated IDPs (r = 0.2994, p = 0.1451, Fig 5C).

### Expansion of HuAd5 hexon-specific CD4⁺ IFN-γ⁺ TNFα⁺ cells in high ChAdOx1 nCoV-19 spike responder IDPs

Having established higher HuAd5 hexon-specific T cells in ChAdOx1 nCoV-19 vaccinated IDPs and a positive association with SARS-CoV-2 spike T cell responses, to further define the functional phenotypes of HuAd5 hexon T cells that may be important in this association, we performed multicolour flow cytometry to assess functional markers of HuAd5 hexon-specific T cells. We then compared the results between high and low SARS-CoV-2 spike-specific T cell responders. Representative plots of the gating strategy are shown in Supplementary Fig 2A. HuAd5 hexon peptide stimulation induced higher frequencies of CD4^+^ IFN-γ^+^ TNFα^+^ T cells in high (SARS-CoV-2 spike) responders compared to low responders (p=0.0002, Fig 6A). Although CD8^+^ IFN-γ^+^ TNFα^+^ T cell frequencies were expanded in some of the high responders, this did not reach significance (p = 0.0505, Supplementary Fig 2A). Notably, both high and low spike responders had similar frequencies of reactive T cells to a mixture of pools of MHC class I and II positive control peptides derived from commonly encountered viruses (Fig 6B). The frequency of HuAd5 hexon-specific CD4^+^ IFN-γ^+^ TNFα^+^ cells correlated strongly with the frequency of SARS-CoV-2 spike-specific T cells (Spearman r = 0.628, p < 0.0001, Fig 6C). As shown in the correlation matrix of the different HuAd5 hexon-specific T cell phenotypes and SARS-CoV-2 spike-specific T cells (Supplementary Fig 2B), CD8^+^ IFN-γ^+^TNFα^+^ cells also had a significant positive correlation with SARS-CoV-2 spike-specific T cells (Spearman r = 0.3980, p = 0.0008, Fig 6D). There was no significant correlation between the different T cell phenotype responses to the control peptides and spike response (Supplementary Fig 2C). We also performed a new analysis of our previously published data on the T cell response to a different pool of positive control peptides^17^ and the results also showed no difference in HuAd5 hexon-specific T cell response between high and low spike responders (p = 0.9723, Supplementary Fig 2D)

**Figure 6:**
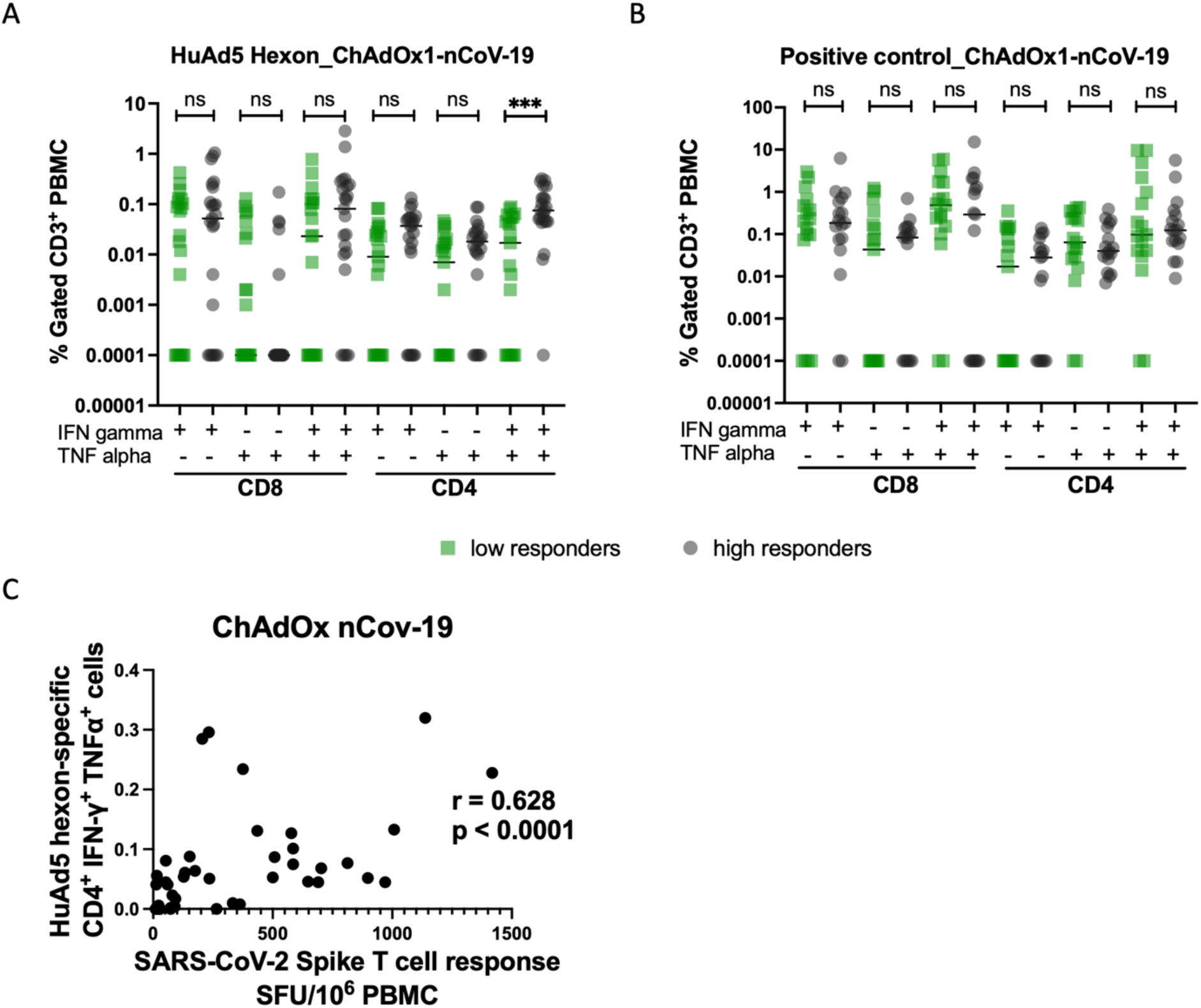
Functional Phenotype of HuAd5 hexon-specific T cells in ChAdOx1 nCoV-19 vaccinated IDPs. PBMC samples from ChAdOx1 nCoV-19 vaccinated IDPs were tested by intracellular cytokine staining and flow cytometry for hexon-specific and positive control-specific T cell subtypes. **A)** Comparison of hexon-specific, cytokine-producing T cell subtypes between high (n = 23) and low (n = 21) spike responders. **B)** Comparison of positive control peptide pool reactive T cell subtypes between high (n = 17) and low (n = 17) reponders. **C)** Correlation between HuAd5 hexon-specific CD4^+^ IFN-γ^+^ TNFα^+^ cells and SARS-CoV-2 spike-specific T-cells. Positive control = CMV, EBV, RSV, IAV and HPIV. Group differences were determined by non-parametric Mann Whitney test: *** is p < 0.001, ns = not significant. Values zero or below were assigned a value of 0.0001 for visualization only. Spearman’s r and p values are shown within the correlation graph.

## DISCUSSION

In this study, we assessed HuAd5 immunity in ChAdOx1 nCoV-19 vaccinated individuals and investigated the mechanisms underlying enhanced ChAdOx1 nCoV-19-induced SARS-CoV-2 spike-reactive T cells and their relationship to pre-existing human adenovirus 5 (HuAd5) immune memory in cohorts of immunodeficient patients (IDPs) and healthcare workers (HCWs). We found that HuAd5 hexon-binding IgG titres were increased in ChAdOx1 nCoV-19 vaccine recipients and higher titres were associated with lower SARS-CoV-2 spike-specific T cell responses in both HCWs and IDPs. Similarly, HuAd5 neutralisation titres was mildly inversely associated with SARS-CoV-2 spike-specific T cell responses in ChAdOx1 nCoV-19 vaccinated IDPs, but titres were similar between cohorts or vaccine groups. We also observed that ChAdOx1 nCoV-19 vaccination significantly increased HuAd5-specific T cell responses. Further immune profiling showed significantly higher frequencies of HuAd5-reactive T cells, particularly CD4⁺ IFN-γ⁺ TNFα⁺ cells, in IDPs with strong spike-specific responses, with a robust positive correlation between these responses. Together, these findings support a model in which cross-reactive adenovirus-specific T cells contribute to bystander enhancement of adenovirus vector-based vaccine-induced cellular immunity, whereas anti-vector antibodies may limit this effect.

Our finding that ChAdOx1 nCoV-19 recipients had higher HuAd5 hexon-binding IgG titres than BNT162b2 recipients in both HCWs and IDPs suggests induction of cross-reactive antibody responses to the vector. HuAd5 neutralisation titres, however, did not show any difference between cohorts or between vaccine groups. Although differences in sensitivity of the assay may be a contributing factor, this discrepancy may be more reflective of differences in immunogenicity and function of the adenoviral structural proteins. The hexon protein is the most abundant and immunogenic capsid component, and may preferentially induce binding antibodies even in the context of weak cross-reactivity ^35^. In contrast, viral entry into lung epithelial cells (simulated in our neutralisation assay) is primarily mediated by fibre and penton base proteins ^36,37^, which are more relevant to neutralizing activity. It is therefore plausible that long-standing, high-affinity neutralizing antibodies from prior HuAd5 exposure dominate neutralization responses, masking any incremental effect of cross-reactive responses induced by ChAdOx1. This may explain our previous findings of lower neutralizing titres against the novel SARS-CoV-2 antigen in these IDPs compared to HCWs^38,39,17^, while HuAd5 neutralizing titres in the current study remained relatively preserved, likely due to repeated natural exposure. Furthermore, in IDPs receiving IgG replacement therapy, the trend toward higher HuAd5 neutralising titres likely reflect passive transfer of durable antibodies, contrasting with their reduced capacity to generate novel antigen-specific responses ^17^. Also, the closely clustered neutralisation titres within this group reflects the fact that the IgG replacement was from a standardised pool from donors, whereas the wide spread in the other cohorts and sub-cohorts reflect diverse immunological history of exposure to adenoviruses.

Higher frequency of HuAd5-specific T cells following ChAdOx1 nCoV-19 compared to BNT162b2 vaccination as we observed in HCWs after first vaccination (Supplementary Fig 1B) and in the IDPs cohort (Fig 2A) supports the concept of cross-recognition between human adenoviruses like HuAd5 and the ChAdY25-derived vaccine vector. We selected HuAd5 as a representative adenovirus because of its high seroprevalence and >70% amino acid similarity to ChAdY25 in key structural proteins, including hexon and penton. Similar cross-reactivity between human and non-human adenoviruses has been described by multiple studies ^40,41,15,12,42^. Gardner et al. (2024) demonstrated early lymphocyte activation following ChAdOx1 stimulation that was absent with BNT162b2, suggesting activation of pre-existing cross-reactive T cells rather than a primary response^12^. More recently, Mukhopadhyay and colleagues demonstrated that ChAdOx1 nCoV-19 vaccination of healthy donors resulted in significantly higher HuAd5 hexon- and ChAdY25 hexon-specific T cells compared to controls^42^. The ability of ChAdOx1 vector to induce cross-reactive T cell responses is an important consideration for furture therapeutic uses of ChAdOx1 and adenovirus vectors especially in people with prior vaccination history.

Our finding that IDPs with stronger SARS-CoV-2 spike-specific T cell responses exhibited higher frequencies of HuAd5-reactive CD4⁺ IFN-γ⁺ TNFα⁺ T cells supports a role for adenovirus-specific CD4⁺ T cells in mediating bystander enhancement of vaccine-induced responses. Importantly, both high and low responders exhibited comparable responses to MHC class I and II-restricted positive control peptide pools from two independent experiments, which supports the specificity of this association, rather than generalized differences in T cell responsiveness. Although enhanced T cell responses following ChAdOx1 vaccination have been attributed to efficient transduction of innate immune cells and sustained antigen presentation,^18,29^ our findings support a perhaps complementary mechanism in which cross-reactive adenovirus-specific T cells act as bystander modulators, amplifying antigen-specific responses following ChAdOx1 vaccination, particularly in immunodeficient individuals.

Importantly, we did find that patients with higher anti-HuAd5 hexon IgG binding titres had lower T cell responses to ChAdOx1 nCoV-19 vaccination and this was the case in both HCWs and IDPs. An inverse correlation was found for HuAd5 neutralization titres and ChAdOx1 nCoV-19 induced spike T cell responses in IDPs and HCWs. This agrees with previous report of higher antibody titres and durability of anti-SARS-CoV-2 spike antibodies following ChAdOx1-n CoV-19 vaccination in individuals without prior human adenovirus antibodies compared to those with pre-existing antibodies^43^. Also, in a ChAd63-based malaria vaccine study, baseline ChAd63 hexon-specific T cell responses, presumed to reflect cross-reactive immunity generated by prior exposure to human adenoviruses, were inversely associated with post-vaccination vaccine-specific antibody responses^44^. Together with our findings, these reports support the concept that naturally acquired immunity to human adenoviruses can influence immune responses to heterologous chimpanzee adenoviral vectors despite limited direct exposure to the vector itself. Our results further suggest that vector cross-reactive antibodies may modulate vaccine-induced T cell responses. Our findings support a mechanism where, in IDPs receiving vector-encoded vaccines, deficiency in anti-vector cross-reactive antibodies allows for more efficient recall of vector-specific T cells leading to expansion and increased cytokine production. This in-turn promotes enhanced vaccine-specific T cell responses.

There are limitations in this study. Firstly, the observational nature of the study precludes definitive conclusion that HuAd5-specific T cells directly mediate enhancement of spike-specific responses and that HuAd5 antibodies inhibit ChAdOx1 nCoV-19 vaccine response. As a retrospective clinical study, it is subject to confounding variables and limited ability to control for demographic differences, particularly given the relatively small sample sizes in some groups. Also, baseline adenovirus immunity was not measured in IDPs and cross-reactivity was inferred from higher HuAd5-specific responses following ChAdOx1 nCoV-19 vaccination rather than being directly measured. In addition, our analysis was restricted to HuAd5 hexon-specific responses; responses to other adenoviral proteins or to additional prevalent adenovirus serotypes were not assessed and may also influence vaccine responses. For example, adenovirus type E and ChAdY25 share even greater amino acid homology of up to 90% in the major structural proteins^45^ and may therefore show greater immune cross-reactivity. Also, T cell immunodominance vary between structural proteins. Lastly, the IDPs are a heterogenous group of primary and secondary immunodeficient patients of various ages and understanding the relevance of these results in different patient subgroups may help best inform targeted vaccination strategies.

Despite these limitations, our findings provide evidence that cross-reactive adenovirus immunity can modulate responses to adenoviral vector vaccines, particularly in antibody-deficient populations. These findings have potential implications for vaccine design and optimization. For example, therapies or protocols could be tailored to suppress vector-specific or cross-reactive antibodies while preserving T cell responses especially in cases where T cell-specific responses are most desirable. Safe vector-based vaccines can be developed for patient groups with predominant antibody deficiency. Given the widespread exposure to adenoviral vectors during the COVID-19 vaccination campaigns, more studies should further investigate the role of cross-reactive T cells and antibodies across a broader range of adenoviral antigens and host immune contexts to better define their impact on vaccine efficacy.

## MATERIALS AND METHODS

### Ethics Statement

This study was approved by the Research Ethics Committee Wales, IRAS: 96194 12/WA/0148. Amendment 5. Written, informed consent was provided to all participants prior to enrolment in the study.

### Human Adenovirus 5-GFP (HuAd5-GFP)

HuAd5-GFP is an E1/E3-deficient adenoviral vector of serotype 5 and was generated as previously described ^46^.

### Cell Line and Culture

A human lung carcinoma cell line, A549 (ATCC, CCl-185) was cultured and passaged in Dulbecco’s modified Eagle medium (DMEM) supplemented with 10 % fetal calf serum (FCS), Gibco, A5670701), 100 U penicillin/ml, 100 mg streptomycin/ml (Thermo Fisher, 15070063). Cells were cultured to 70 – 90 % confluency at 37 ° C, 5 % CO_2_. For passage, cells were detached with trypsin and EDTA treatment for 3 min, washed and resuspended in cell culture medium. Live cells were counted using the trypan blue exclusion method.

### Study Cohorts

Participants in this study are as described in our previous publication^17^. Briefly, SARS-CoV-2 infection-naïve immunodeficient out-patients (IDPs) with diagnosed primary or secondary immune deficiency under the Respiratory Immunology Service, Royal Papworth Hospital (RPH) were recruited between March and July 2021. Immune diagnosis and treatment with immunoglobulin replacement therapy (IgGRx) were recorded. SARS-CoV-2 infection-naïve healthcare workers (HCWs) at the Royal Papworth Hospital were recruited from the Humoral Immune Correlates for COVID-19 (HICC) study as healthy controls. Both IDPs and HCWs subsequently received double homologous doses of either ChAdOx1 n-CoV-19 or BNT162b2 as described in our previous publication. Study participants having SARS-CoV-2 spike T cell responses below the limit of detection, as previously determined^17^, were not tested for HuAd5 hexon-specific T cell responses and excluded from T cell analyses. The ages and other demographic information of study participants are as previously described^17^.

### Sample Processing

Whole blood samples were collected and processed for serum and PBMC as previously described ^17^.

### IFN-γ ELISpot

*Ex vivo* ELISpot assays were performed using the Human IFN-γ Single-Colour Enzymatic ELISPOT Assay kit (ImmunoSpot®) according to the manufacturer’s protocol. Briefly, PVDF membranes were coated with anti-IFN-γ capture antibody overnight and then washed once with PBS. 100 µL of peptide solution in CTL-Test^TM^ medium was plated per well at a final peptide concentration of 1 µg/ml. Peptides were 15mers derived from HuAd5 hexon (PepMix™ Human Adenovirus 5, JPT, PM-HADV5-L3-1). Cell Stimulation Cocktail (containing Phorbol 12-myrstate 13-acetate (PMA) and ionomycin, eBioscience) was used as an experiment positive control at 1:500 dilution. As a negative control, an equimolar concentration of DMSO in CTL-Test^TM^ medium was used. PBMCs were added at 2 × 10^5^ cells/well in 100 µl in CTL-Test^TM^ medium and the plate immediately transferred to a humidified incubator at 37 ° C, 5 % CO_2_ for 24 h. Cells were then decanted, and the detection antibody was added. Following this, the membrane-bound cytokine was visualized by an enzymatic reaction. Plates were dried, scanned and spots were analysed using the ImmunoSpot® S6 Ultra-V analyser (CTL, software: CTL Switchboard 2.7.2 x64). For all wells, the number of spot-forming units (SFU) were determined using SmartCount^TM^ with gates adjusted to positive and negative controls per plate. Tests and controls were carried out in duplicate for each sample. Counts per sample were obtained by subtracting the mean of background SFU (DMSO control) from the mean of peptide-stimulated SFU then expressed as SFU/million PBMCs. Zero or negative values were assigned a value of 1.

### Intracellular Cytokine Staining and Flow Cytometry

Cells were incubated at 37 ° C, 5 % CO_2_ in complete medium (RPMI 1640-glutamax containing 1 mM sodium pyruvate, 100 U penicillin/ml, 100 μg streptomycin/ml, 50 uM β-mercaptoethanol and 10 % FCS) containing HuAd5 hexon peptides at a concentration of 2 µg/ml. As a positive control, a mixture of CERI-MHC Class I Control Peptide pool made up of Cytomegalovirus (CMV), Epstein Barr virus (EBV), Respiratory Syncytia virus (RSV), Influenza A virus (IAV) peptides (ImmunoSpot®, CTL-CERI-300)^47^ and CPI MHC II Positive Control Solution made up of CMV, IAV, Human Parainfluenza virus (HPIV) peptides (ImmunoSpot®, CTL-CPI-001)^47^ was used at 1x concentration to stimulate cells. The volume of medium per well of 96 well U-bottom plates (Greiner) was 100 μl. 2 h into the stimulation, 50 μl of complete medium containing 3x protein transport inhibitor cocktail (PTI) (eBioscience, 00-4980-93) was added (to achieve 1x concentration of PTI in cell culture) and cells were incubated for another 12 h. Cells were pelleted by centrifugation at 330 x ***g*** for 3 min and supernatant was removed. This centrifugation speed and time was used for the subsequent wash and pelleting steps. Cells were washed with wash buffer (1X PBS supplemented with 2.5 % concentrated FBS and 2 mM EDTA) and stained for viability and the T cell markers CD3, CD4 and CD8. After 30 min incubation at 4 ° C, cells were washed twice with wash buffer, fixed using the Cytofix/Cytoperm™ Fixation/Permeabilization Kit (BD Biosciences) for 10 min at room temperature and washed with wash buffer. For the intracellular detection of IFN-γ and TNF-α, cells were incubated for 30 min with the fluorochrome conjugated antibodies (Table 2.4) in 1X BD Perm/Wash buffer. Cells were washed twice in Perm/Wash buffer, resuspended in wash buffer and analysed using the Attune Nxt flow cytometer (Invitrogen. Software 6.2.1).

### Flow Cytometry panel

| Laser | Filter | Product | Antigen | Fluorochrome | Source | Cat. No. |
| --- | --- | --- | --- | --- | --- | --- |
| Blue:<br>488 | 530/30 | CD4 Monoclonal Antibody (OKT4) | CD4 | FITC | Thermofisher | 11-0048-42 |
| Yellow:<br>561 | 620/15 | TNF alpha Monoclonal Antibody (MAb11) | TNF-a | PE efluor610 | Thermofisher | 61-7349-42 |
|  | 695/40 | CD3 Monoclonal Antibody (SK7) | CD3 | PE Cy5.5 | Thermofisher | 35-0036-42 |
| Red:<br>637 | 670/14 | IFN gamma Monoclonal Antibody (B27) | IFN $\gamma$ | APC | Thermofisher | MHCIFG05 |
|  | 720/30 | APC-R700 Mouse Anti-Human CD8 | CD8 | APC R-700 | BD Bioscience | 565192 |
|  | 780/60 | Fixable Viability Dye | Live/dead | eFluor 780 | eBioscience | 65-0865-14 |

### Luminex Multiplex Assay

HuAd5 hexon protein (Native Antigen Company, SKU: AH01) was coupled to Luminex beads using methods originally described in the Luminex (xMAP) Cookbook (4th Edition) and as in Castillo-Olivares et al. (2021) and Xiong et al. (2020). Briefly, 1.25 × 10^7^ superparamagnetic MagPlex ®-c Microsphere beads (Bio-Rad) were activated with 1-ethyl-3-(3-dimethylaminopropyl) carbodiimide hydrochloride (Thermo Fisher Scientific, E2247) in the presence of N-hydroxysuccinimide (Thermo Fisher Scientific, 24500) in 100 mM MES buffer (Thermo Fisher Scientific, J61587.AP), pH 6.0. After washing with Dulbecco’s PBS (DPBS), pH 7.4 (without Mg^2+^/Ca^2+^) (Gibco, 14040133), 50 μg of the protein to be coupled to beads were incubated with the beads in DPBS, pH 7.4, for 2 h on a rotator at room temperature. Coupled beads were then washed with DPBS and stored in storage buffer (DPBS, 1 % BSA, and 0.05 % w/v NaN_3_) at +4 ° C.

At a final concentration of 30 beads per µL in an assay volume of 50 µL, beads were incubated with serum diluted 1:500 with assay buffer (DPBS (without Mg^2+^/Ca^2+^), 1 % BSA (Sigma, A9418-50G), 0.8 % Poly Vinyl Pyrrolidone (Sigma, 9003-39-8), 0.5 % Poly Vinyl Alcohol (Sigma, 9002-89-5), 0.05 % w/v NaN_3_ (Sigma, 26628-22-8)) and incubated for 1 h at 37 ° C on an orbital shaker at 500 rpm in the dark. The beads were washed three times with 100 µL wash buffer (PBST: DPBS without Mg^2+^/Ca^2+^ and 0.05 % v/v Tween-20 (Sigma, 9005-64-5)). A magnetic holder (Bio-Rad, 1614916) was used to prevent loss of beads during washing. Anti-Human IgG Fc Specific antibody PE conjugated detection antibody (Leinco Technologies, Inc. I-127) was diluted to 2 µg/mL in assay buffer and 50 µL was added to all wells. Plates were incubated in the dark at 37 ° C on an orbital shaker for 30 min. After washing three times as before, the beads were resuspended in 100 µL of wash buffer and analysed on a Bio-Rad Bio-Plex 200. The data was captured with the Bio-Plex Manager Software and analysed using Microsoft Excel and GraphPad Prism 9. The data are reported as Mean Fluorescence Intensity (MFI) less background, with background being a serum sample-free buffer control.

### Human Adenovirus 5-GFP (HuAd5-GFP) neutralisation assay

Replication-deficient HuAd5 with a E1/E3 deletion, encoding GFP was used in neutralisation assays to analyse the inhibitory effect of diluted serum on adenovirus transduction of A549 cells. Sera were tested in duplicate in 5 point 5-fold serial dilutions (1:10, 1:50, 1:250, 1:1250, 1:6250). Diluted sera were incubated with 2 × 10^5^ infectious units of HuAd5-GFP per well for 1 h. Cells were added at 2 × 10^4^ cells/well to achieve a multiplicity of infection (moi) of 10. The set up was then incubated for 24 h. As controls, cells with HuAd5-GFP but no serum were used to represent 0 % neutralisation, while cells only with no virus or serum represent 100 % neutralisation or background GFP expression. At the end of the incubation period, cells were inspected using fluorescence microscopy. Supernatants were then discarded leaving the adherent cells which were then detached using trypsin and EDTA treatment for 3 min and washed once in Hank’s solution. Cells were resuspended in Hank’s solution and analysed by flow cytometry using the Attune Nxt flow cytometer for GFP production. The mean of percentage gated GFP^+^ cells was the assay readout and was obtained for each serum dilution per sample. Results were then normalized to controls and plotted to obtain area under the curve (AUC).

### Result Analysis based on Previously Published Data

HuAd5 T cell and antibody responses in this study were analysed based on our previously published data on SARS-CoV-2 T cell responses in this same cohort ^17^. To allow group comparisons, SARS-CoV-2 spike-specific T cell responders were divided into two groups based on a cut off of 200 spot forming units from the ELISpost assay. Samples with ≥ 200 spots (per million PBMCs) were considered high responders while samples with ≥ 8 and < 199 spots were considered low responders. This cut off point also divided the cohort into approximately 2 equal halves. 8 SFU was our limit of detection based on our pre-determined cut-off.

### Statistical Analysis

Statistical analyses were performed using Graphpad Prism Software (GraphPad Prism Software, CA), version 10. Data were analysed to determine statistical differences using the non-parametric Mann-Whitney test. Correlation was determined using Spearman’s correlation. Graphs were produced using Graphpad Prism 10.

## ACKNOWLEDGEMENTS

This work was supported by the NIHR/UKRI grant (COV0170) Humoral Immune Correlates of COVID-19 (HICC), NIHR/UK-HSA (MR/W02067X/1) SIREN study, Royal Papworth Hospital NHS Foundation Trust Charity and Cambridge Commonwealth, European & International Trust (Cambridge Trust) in association with Cambridge-Africa. We thank the Royal Papworth Hospital Foundation Trust COVID-19 Research and Clinical teams including the Papworth Trials Unit Collaboration (PTUC), and the NIHR Study Support Service LCRN Eastern Core Team for supporting recruitment to this study. We thank the healthcare workers and Outpatients who participated in this study. We thank Jessica Gronlund and Helen Gronlund who assisted with coordinating sample collection and Dr Sofiya Fedosyuk for assisting with manuscript review.

## AUTHOR CONTRIBUTION

E.T.A. conceived the main ideas of the study and designed the study. H.B and J.L.H. acquired funding. B.B, H.B and J.L.H. supervised the study. B.A and R.W provided and validated key reagents. J.C.O served as study coordinator. E.T.A, A.N, A.CY.C processed samples and performed the assays. E.T.A wrote the manuscript with contributions from G.W.C and B.B. Data analysis and visualization were performed by E.T.A. The following authors reviewed the manuscript and provided critical feedback: G.W.C, B.B, H.B, J.L.H. All authors contributed substantially to the discussion of the content and/or edited the manuscript before submission.

## ETHICAL DECLARATIONS

### Data availability

Further information and requests should be directed to and will be fulfilled by the lead contacts, E.T.A and J.L.H.

### License information

For the purpose of Open Access, the author has applied a CC BY public copyright licence to any Author Accepted Manuscript (AAM) version arising from this submission.

## SUPPLEMENTARY FIGURES

**Supplementary Figure 1:**
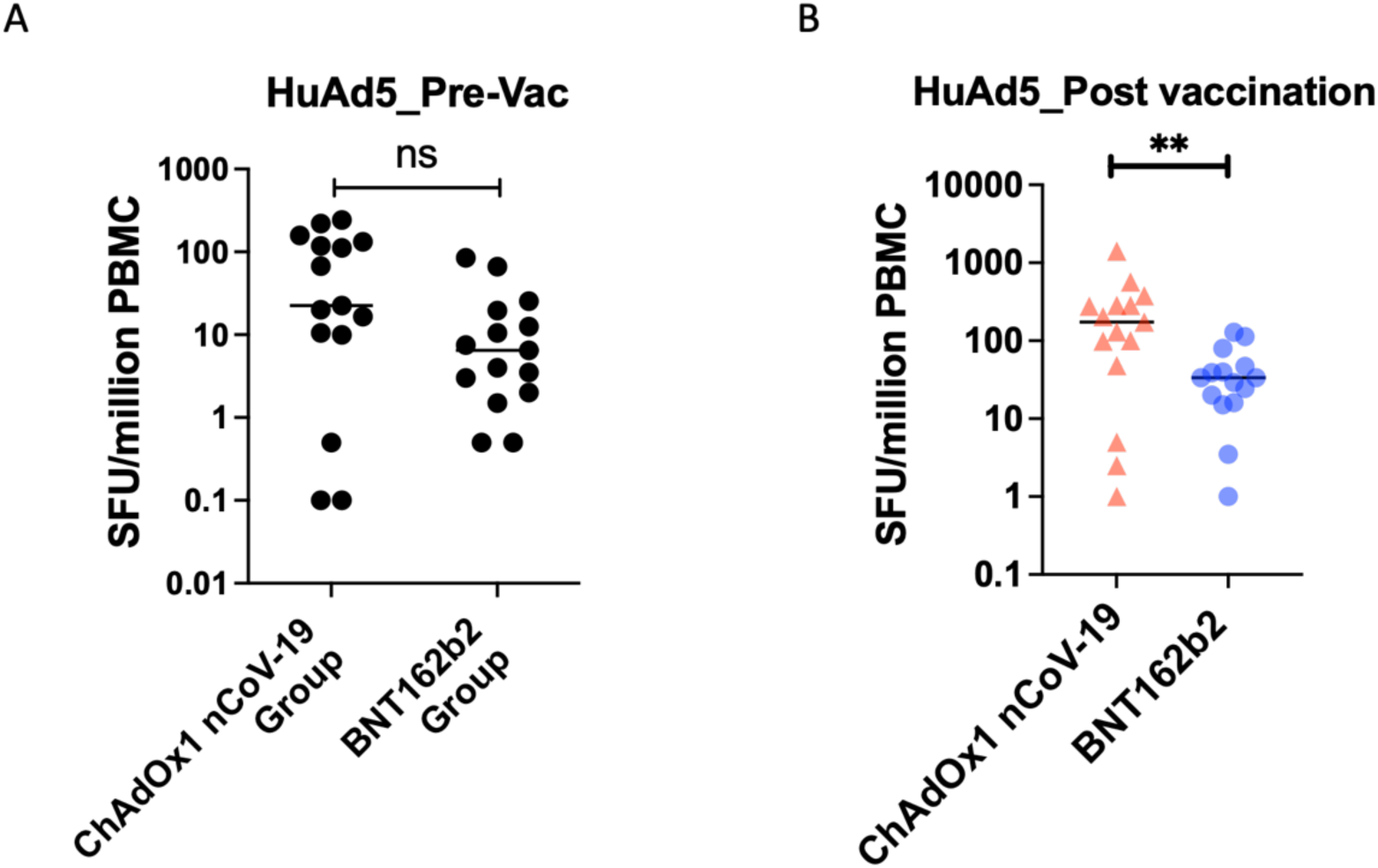
HuAd5 Hexon-specific T Cell Responses in HCWs. A & B) Fifteen paired PBMC samples from HCW pre-vaccination and post-first dose of COVID-19 vaccines (ChAdOx1-nCoV-19 or BNT162b2) were stimulated with HuAd5 hexon protein peptides and T cell responses were assessed using the ELISpot assay. Comparison of HuAd5 hexon-specific T cell responses between vaccine groups **A)** pre-vaccination and **B)** post first vaccination. Data points are mean of duplicate wells minus negative controls. Significant differences were determined by non-parametric Mann Whitney test: ** is p < 0.01, ns = not significant.

**Supplementary Figure 2:**
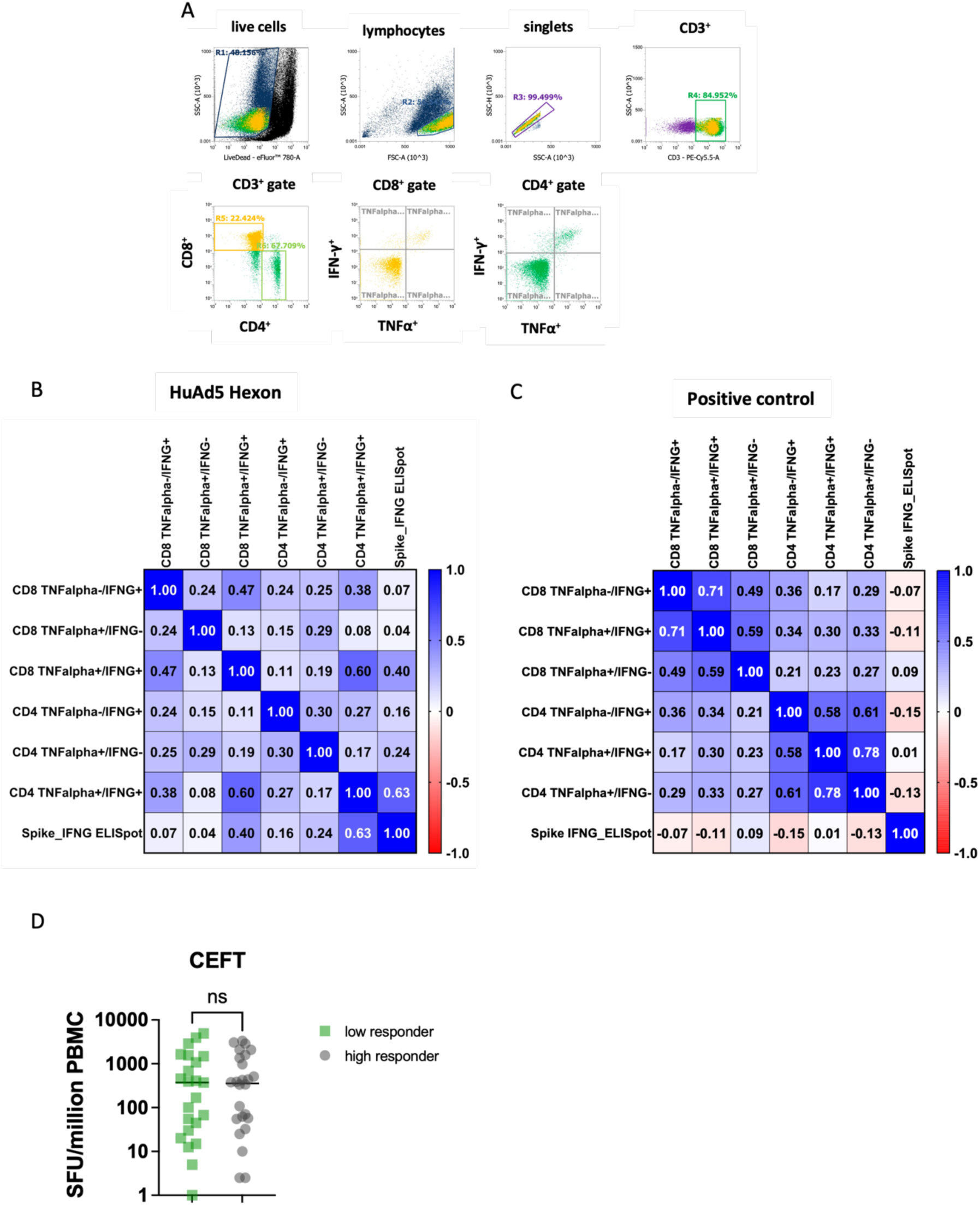
Functional Phenotype of HuAd5 hexon-specific T cells in ChAdOx1 nCoV-19 vaccinated IDPs. PBMC samples from ChAdOx1 nCoV-19 vaccinated IDPs were tested by intracellular cytokine staining and flow cytometry for hexon-specific and positive control-specific T cell subtypes. **A)** Representative flow cytometry dot plots indicating gating strategy. **B)** correlation matrix of hexon-specific T cell phenotypes and SARS-CoV-2 spike T-cell response. **C)** correlation matrix of positive control-specific T cell phenotypes and SARS-CoV-2 spike T-cell response. Positive control = pool of peptides derived from Cytomegalovirus, Epstein Barr virus, Respiratory Syncytia virus, Influenza A virus, Parainfluenza. **D)** New analysis of previously published data^17^ comparing CEFT-specific T cells between high (n = 26) and low (n = 24) SARS-CoV-2 responders within ChAdOx1 nCoV-19 vaccinated IDPs. CEFT = CMV, EBV, Flu, Tetanus.

## REFERENCES

1. Draper, S. J. & Heeney, J. L. Viruses as vaccine vectors for infectious diseases and cancer. Nat Rev Microbiol 8, 62–73 (2010).

2. McCann, N., O’Connor, D., Lambe, T. & Pollard, A. J. Viral vector vaccines. Curr Opin Immunol 77, 102210 (2022).

3. Voysey, M. et al. Safety and efficacy of the ChAdOx1 nCoV-19 vaccine (AZD1222) against SARS-CoV-2: an interim analysis of four randomised controlled trials in Brazil, South Africa, and the UK. The Lancet 397, 99–111 (2021).

4. Mennechet, F. J. D. et al. A review of 65 years of human adenovirus seroprevalence. Expert Rev Vaccines 18, 597–613 (2019).

5. Barouch, D. H. et al. Immunogenicity of Recombinant Adenovirus Serotype 35 Vaccine in the Presence of Pre-Existing Anti-Ad5 Immunity1. The Journal of Immunology 172, 6290–6297 (2004).

6. Li, J.-X. et al. Immunity duration of a recombinant adenovirus type-5 vector-based Ebola vaccine and a homologous prime-boost immunisation in healthy adults in China: final report of a randomised, double-blind, placebo-controlled, phase 1 trial. Lancet Glob Health 5, e324–e334 (2017).

7. Pine, S. O. et al. Pre-Existing Adenovirus Immunity Modifies a Complex Mixed Th1 and Th2 Cytokine Response to an Ad5/HIV-1 Vaccine Candidate in Humans. PLoS One 6, e18526 (2011).

8. Zhu, F.-C. et al. Safety, tolerability, and immunogenicity of a recombinant adenovirus type-5 vectored COVID-19 vaccine: a dose-escalation, open-label, non-randomised, first-in-human trial. Lancet 395, 1845–1854 (2020).

9. Sadoff, J. et al. Interim Results of a Phase 1-2a Trial of Ad26.COV2.S Covid-19 Vaccine. N Engl J Med 384, 1824–1835 (2021).

10. Tapia, M. D. et al. Use of ChAd3-EBO-Z Ebola virus vaccine in Malian and US adults, and boosting of Malian adults with MVA-BN-Filo: a phase 1, single-blind, randomised trial, a phase 1b, open-label and double-blind, dose-escalation trial, and a nested, randomised, double-blind, placebo-controlled trial. Lancet Infect Dis 16, 31–42 (2016).

11. Dicks, M. D. J. et al. A Novel Chimpanzee Adenovirus Vector with Low Human Seroprevalence: Improved Systems for Vector Derivation and Comparative Immunogenicity. PLOS ONE 7, e40385 (2012).

12. Gardner, J. et al. Identification of cross reactive T cell responses in adenovirus based COVID 19 vaccines. npj Vaccines 9, 1–12 (2024).

13. Barouch, D. H. et al. Characterization of humoral and cellular immune responses elicited by a recombinant adenovirus serotype 26 HIV-1 Env vaccine in healthy adults (IPCAVD 001). J Infect Dis 207, 248–256 (2013).

14. Heemskerk, B. et al. Extensive Cross-Reactivity of CD4+ Adenovirus-Specific T Cells: Implications for Immunotherapy and Gene Therapy. J Virol 77, 6562–6566 (2003).

15. Iampietro, M. J. et al. Immunogenicity and Cross-Reactivity of Rhesus Adenoviral Vectors. J Virol 92, e00159–18 (2018).

16. Righi, E. et al. A Review of Vaccinations in Adult Patients with Secondary Immunodeficiency. Infect Dis Ther 10, 637–661 (2021).

17. Aguinam, E. T. et al. Differential T-cell and antibody responses induced by mRNA versus adenoviral vectored COVID-19 vaccines in patients with immunodeficiencies. J Allergy Clin Immunol Glob 2, 100091 (2023).

18. Parry, H. et al. Immunogenicity of single vaccination with BNT162b2 or ChAdOx1 nCoV-19 at 5–6 weeks post vaccine in participants aged 80 years or older: an exploratory analysis. The Lancet Healthy Longevity 2, e554–e560 (2021).

19. Parry, H. et al. Differential immunogenicity of BNT162b2 or ChAdOx1 vaccines after extended-interval homologous dual vaccination in older people. Immunity & Ageing 18, 34 (2021).

20. Prendecki, M. et al. Humoral and T-cell responses to SARS-CoV-2 vaccination in patients receiving immunosuppression. Ann Rheum Dis 80, 1322–1329 (2021).

21. Saleem, B., Ross, R. L., Duquenne, L., Hughes, P. & Emery, P. COVID-19 vaccine-induced T-cell responses in patients with rheumatoid arthritis: preferential induction by ChAdOx1. The Lancet Rheumatology 4, e171–e172 (2022).

22. Kim, T.-S. & Shin, E.-C. The activation of bystander CD8+ T cells and their roles in viral infection. Exp Mol Med 51, 1–9 (2019).

23. Lee, H.-G., Cho, M.-J. & Choi, J.-M. Bystander CD4+ T cells: crossroads between innate and adaptive immunity. Exp Mol Med 52, 1255–1263 (2020).

24. Shim, C.-H., Cho, S., Shin, Y.-M. & Choi, J.-M. Emerging role of bystander T cell activation in autoimmune diseases. BMB Rep 55, 57–64 (2022).

25. Di Genova, G., Savelyeva, N., Suchacki, A., Thirdborough, S. M. & Stevenson, F. K. Bystander stimulation of activated CD4+ T cells of unrelated specificity following a booster vaccination with tetanus toxoid. European Journal of Immunology 40, 976–985 (2010).

26. van Aalst, S., Ludwig, I. S., van der Zee, R., van Eden, W. & Broere, F. Bystander activation of irrelevant CD4+ T cells following antigen-specific vaccination occurs in the presence and absence of adjuvant. PLoS One 12, e0177365 (2017).

27. Maurice, N. J., McElrath, M. J., Andersen-Nissen, E., Frahm, N. & Prlic, M. CXCR3 enables recruitment and site-specific bystander activation of memory CD8+ T cells. Nat Commun 10, 4987 (2019).

28. Chapter 9 - T Cell Development, Activation and Effector Functions. in Primer to the Immune Response (Second *Edition)* (eds Mak, T. W., Saunders, M. E. & Jett, B. D.) 197–226 (Academic Cell, Boston, 2014). doi:10.1016/B978-0-12-385245-8.00009-1.

29. Provine, N. M. & Klenerman, P. Adenovirus vector and mRNA vaccines: Mechanisms regulating their immunogenicity. Eur J Immunol 10.1002/eji.202250022 (2022) doi:10.1002/eji.202250022.

30. Yang, T. C., Dayball, K., Wan, Y. H. & Bramson, J. Detailed Analysis of the CD8+ T-Cell Response following Adenovirus Vaccination. J Virol 77, 13407–13411 (2003).

31. Yang, T.-C. et al. The CD8+ T Cell Population Elicited by Recombinant Adenovirus Displays a Novel Partially Exhausted Phenotype Associated with Prolonged Antigen Presentation That Nonetheless Provides Long-Term Immunity1. The Journal of Immunology 176, 200–210 (2006).

32. Li, C. et al. Mechanisms of innate and adaptive immunity to the Pfizer-BioNTech BNT162b2 vaccine. Nat Immunol 23, 543–555 (2022).

33. Basner-Tschakarjan, E. et al. Adenovirus efficiently transduces plasmacytoid dendritic cells resulting in TLR9-dependent maturation and IFN-alpha production. J Gene Med 8, 1300–1306 (2006).

34. Arunachalam, P. S. et al. Systems vaccinology of the BNT162b2 mRNA vaccine in humans. Nature 596, 410–416 (2021).

35. Leen, A. M., Bollard, C. M., Myers, G. D. & Rooney, C. M. Adenoviral Infections in Hematopoietic Stem Cell Transplantation. Biology of Blood and Marrow Transplantation 12, 243–251 (2006).

36. Zubieta, C., Schoehn, G., Chroboczek, J. & Cusack, S. The Structure of the Human Adenovirus 2 Penton. Molecular Cell 17, 121–135 (2005).

37. Greber, U. F. & Suomalainen, M. Adenovirus entry: Stability, uncoating, and nuclear import. Mol Microbiol 118, 309–320 (2022).

38. Nadesalingam, A. et al. Paucity and discordance of neutralising antibody responses to SARS-CoV-2 VOCs in vaccinated immunodeficient patients and health-care workers in the UK. The Lancet Microbe 0, (2021).

39. Nadesalingam, A. et al. Vaccination and protective immunity to SARS-CoV-2 omicron variants in people with immunodeficiencies. Lancet Microbe 4, e58–e59 (2022).

40. Saggau, C. et al. The pre-exposure SARS-CoV-2-specific T cell repertoire determines the quality of the immune response to vaccination. Immunity 55, 1924–1939.e5 (2022).

41. Hutnick, N. A., Carnathan, D., Ertl, H. & Betts, M. R. Adenovirus-Specific Human T cells are Pervasive, Polyfunctional, and Cross Reactive. Vaccine 28, 1932–1941 (2010).

42. Mukhopadhyay, R. et al. Adenovirus-Specific T Cells in Adults Are Frequent, Cross-Reactive to Common Childhood Adenovirus Infections and Boosted by Adenovirus-Vectored Vaccines. J Med Virol 97, e70222 (2025).

43. Kim, J. et al. Immunogenicity Differences of the ChAdOx1 nCoV-19 Vaccine According to Pre-Existing Adenovirus Immunity. Vaccines (Basel) 11, 784 (2023).

44. Bliss, C. M. et al. Assessment of novel vaccination regimens using viral vectored liver stage malaria vaccines encoding ME-TRAP. Scientific Reports 8, 3390 (2018).

45. Pm, F., et al. Vaccines based on the replication-deficient simian adenoviral vector ChAdOx1: Standardized template with key considerations for a risk/benefit assessment. PubMed https://pubmed.ncbi.nlm.nih.gov/35715352/ (2022).

46. Ruzsics, Z., Lemnitzer, F. & Thirion, C. Engineering adenovirus genome by bacterial artificial chromosome (BAC) technology. Methods Mol Biol 1089, 143–158 (2014).

47. Lehmann, A. A., Reche, P. A., Zhang, T., Suwansaard, M. & Lehmann, P. V. CERI, CEFX, and CPI: Largely Improved Positive Controls for Testing Antigen-Specific T Cell Function in PBMC Compared to CEF. Cells 10, 248 (2021).

